# An Automated Pipeline for the Identification of Liver Tissue in Ultrasound Video

**DOI:** 10.1101/2024.08.01.24311342

**Authors:** Eloise Ockenden, Simon Mpooya, J. Alison Noble, Goylette F. Chami

## Abstract

Liver diseases are a leading cause of death worldwide, with an estimated 2 million deaths each year. Causes of liver disease are difficult to ascertain, especially in sub-Saharan Africa where there is a high prevalence of infectious diseases such as hepatitis B and schistosomiasis, along with alcohol use. Point-of-care ultrasound often is used in low-resource settings for diagnosis of liver disease due to its portability and low cost. For classification models that can automatically stage liver disease from ultrasound video, the region of interest is liver tissue. A fully-automated pipeline for liver tissue identification in ultrasound video is presented. Ultrasound video data was collected using a low-cost, portable ultrasound machine in rural areas of Uganda. The pipeline first detects the diaphragm in each ultrasound video frame, then segments the diaphragm to ultimately use this segmentation to infer the position of liver tissue in each frame. This pipeline outperforms directly segmenting liver tissue with an intersection over union of 0.83 compared to 0.62. This pipeline also shows improved results with respect to the ease of clinical interpretation and anticipated clinical utility.

## 1 Introduction

Liver diseases are a leading cause of death in sub-Saharan Africa, with commonly recorded causes of hepatitis B and alcohol use [18, 7]. A less recorded, but widely prevalent cause of chronic, severe liver disease in areas of sub-Saharan Africa without access to safe water and adequate sanitation is schistosomiasis. Chronic schistosomiasis infection can cause liver fibrosis, due to immune responses to eggs from intestinal species of this parasitic blood fluke that are trapped in vessels in the portal system [15, 1]. This type of liver fibrosis is called schistosomal periportal fibrosis (PPF). In its early stages, schistosomal PPF is visible at the early segmental branches of the portal system then advances towards the main portal vein, and finally extends across the whole liver [3, 17].

Ultrasound imaging is the most-used, and only recommended method by the World Health Organisation (WHO) for the diagnosis of schistosomal PPF, used due to the high echogenicity of the fibrosis [12]. To reach rural populations affected by schistosomiasis and provide point-of-care services where health systems lack diagnostic capacity, low-cost, portable ultrasound machines are used [5]. It has been shown that the quality of images attained by low-cost, portable ultrasound machines may be lower than the quality obtained from cart-based ultrasound machines [9, 20]. To enable liver tissue identification in this context, deep learning approaches might need to exploit features or anatomy that are known to appear with clearer boundaries.

Automated liver tissue identification from ultrasound video has not been widely explored, especially considering challenges posed in low-resource settings. Segmentation for liver ultrasound has focused on cancer, in particular segmenting focal lesions as a pre-processing step before classification [16]. There has been work on the segmentation of liver tissue in ultrasound images, using data collected in hospital settings. A proposed method has been to locate the liver capsule and use this as a boundary for liver tissue, before cirrhosis staging [14]. However, this approach was used only for ultrasound images that did not image deep into the body, which do not capture enough liver tissue for staging complex diseases that present in both diffuse and focal forms, such as schistosomal PPF. Recently, another method has been proposed, achieving very good quantitative results by using multi-head self attention to segment liver tissue and related structures in ultrasound images [21]. Despite this, there remains no literature on the identification of liver tissue in ultrasound video. Ultrasound video may be a more appropriate data form in low-resource settings. The acquisition of specific views of the liver to produce curated images requires expert knowledge that may not be available. Alternatively, predefined sweep procedures for video acquisition requires less expertise and training. Thus, there remains a need to investigate liver tissue identification using ultrasound video collected in low-resource settings.

In this paper, a pipeline is presented to automatically identify when and where liver tissue appears in ultrasound video, using ultrasound examinations of individuals in a rural, sub-Saharan African context.

## 2 Methods

### 2.1 Context and data

This study was conducted within SchistoTrack, which is a community-based cohort in rural Uganda [2]. Data from an annual follow-up of 3219 participants in 2023 were used. Ultrasound videos were collected from each participant following a predefined protocol conducted by local sonographers who were highly experienced in assessing liver disease. The procedure for acquiring the video used for the development of this pipeline was to start with a good view of the gall bladder with the probe transverse, and then to move the probe to the cephalic position. Each video clip was 5 seconds long, at 20 frames per second (fps). Phillips C5-2 curvilinear transducers were used with the Lumify Application connected to tablets with Android 9 Pie.

Adults who had fasted before the scan and had no ultrasound findings including liver fibrosis, cirrhosis, fatty liver, hepatitis B and other abnormalities, were used for the development of the pipeline. Of the 728 adults who fit these criteria, a random sample of 110 adults were selected, each with one ultrasound video. The participants’ age range was 18-84 years, with a mean (s. d.) of 42.0 (15.6) and 77% (85) were female. Every other frame of each video was labelled with whether or not the diaphragm was visible. Additionally, masks of the diaphragm were created for every other frame of ultrasound video for a randomly selected 25 videos. For diaphragm detection, 90 videos were used for training, 10 for validation, and 10 for testing. For automated diaphragm segmentation, 18 videos were used for training, two for validation, and five for testing.

### 2.2 Diaphragm Detection

As the first step of the pipeline, diaphragm detection was conducted. In B-mode liver ultrasound imaging, the diaphragm is viewed as a echogenic line between the lung and the liver [10]. The diaphragm appears echogenic and clear even in cases where liver tissue as a whole may have an ill-defined boundary. The model used for diaphragm detection was a fine-tuned PulseNet encoder with support vector machines (SVM) as the classification head. The PulseNet encoder used the SonoNet model [4], fine-tuned on training data from the PULSE study [8]. The encoder was trained originally for classification of 13 fetal anatomy views [11], and fine-tuned for diaphragm detection. Predictions were considered in the context of temporal dependence between frames of video. If one isolated frame was predicted to have no diaphragm while neighbouring frames were predicted otherwise, the isolated frame was reclassified as containing the diaphragm. Similarly, if three or fewer consecutive frames were predicted to have a diaphragm, while their surrounding frames were predicted to have no diaphragm, these frames were reclassified to have no diaphragm. This temporal smoothing was based on the appearance of the diaphragm in the training set. The diaphragm never appeared in less than three frames consecutively nor disappeared ever for one isolated frame.

#### Hyperparameters and training

Bayesian hyperparameter tuning was used for the PulseNet encoder. The temperature parameter, used for estimating uncertainty in the dataset, was tuned to a value of 1.25. The learning rate was tuned to a value of 0.022. The model was trained for 100 epochs with early stopping implemented if the validation accuracy stayed within a window of 0.02 for 10 consecutive epochs. The batch size was 32.

### 2.3 Diaphragm Segmentation for Liver Tissue Identification

In order to use the diaphragm as an anchor for inferring where liver tissue appears, it must be segmented. Segmentation masks were propagated across video frames using adaptive memory to exploit the temporal dependence between frames as described elsewhere [22]. The original model was trained on fetal ultrasound video for segmentation of the maternal bladder and placenta, and was fine-tuned here using 20 annotated videos of the diaphragm. During inference, to initialise the model for a video, a segmentation mask of the target object (in this case, the diaphragm) for the first frame of ultrasound video was required. To fully automate the use of this model, a bank of ultrasound images was created from the training set. The bank contained liver views with diverse representations of the diaphragm that had been labelled with masks of the diaphragm. An ultrasound image was selected from this bank that best matched the first frame of ultrasound video, which was identified using the structural similarity index measure (SSIM) [19]. The labelled ultrasound image from the bank of ultrasound images and masks that had the highest SSIM value with the first frame of ultrasound video was used to initialise the model.

Using the diaphragm as an anatomical anchor, the position of liver tissue was identified by finding the end-points of the mask of the diaphragm, and linking these up to the closest points on the left and right edges of the field of view. There were some cases where the diaphragm stretched less than halfway across the field of view. When finding the closest point in Euclidean space, it was often the case that this point was towards the top of the field of view. Therefore, using this point for partitioning created a crude under-estimate of liver tissue by cutting out the lower right side of the field of view. Accounting for the fan shape of the ultrasound field of view, the closest point in Euclidean space was corrected in cases with partial diaphragm observation by moving the point used to create the partition towards the bottom of the field of view, by a factor of 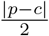, where *p* is the original closest point, and *c* is the midpoint of the right-hand edge of the field of view.

### 2.4 Full Pipeline

The methods described above fit into a fully-automated liver tissue identification pipeline, presented in Figure 1.

**Fig. 1.**
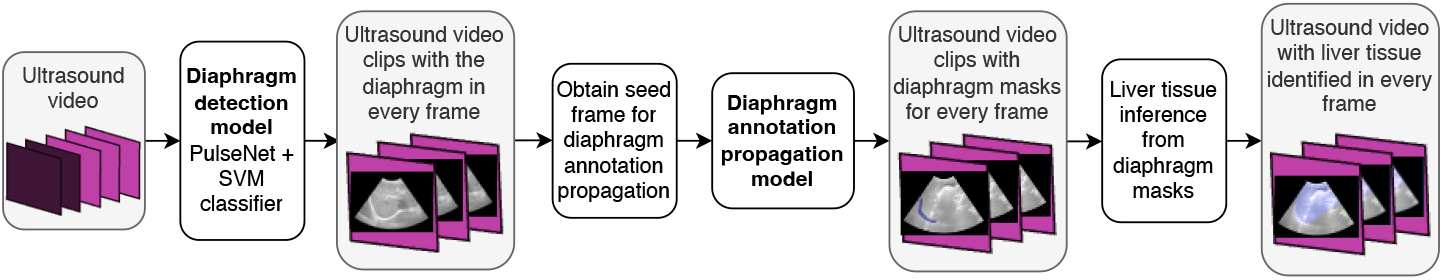
Automated pipeline for liver tissue identification from ultrasound video.

#### Performance metrics and benchmarks

Performance was evaluated using sensitivity, specificity, and F1 score for diaphragm detection. For the final liver tissue identification, intersection over union (IoU) was used as a quantitative evaluation metric, in addition to the investigation of failure cases and clinical utility. For benchmarking diaphragm detection, ResNet-50 with cross entropy (CE) loss and a ResNet-50 using focal loss to account for the class imbalance [13] were used. The ResNet models were pre-trained on the ImageNet dataset [6]. As a benchmark for the full pipeline, the annotation propagation model detailed in [22] was applied without the full pipeline to directly segment liver tissue.

## 3 Experiments and Results

### 3.1 Diaphragm Detection

For diaphragm detection, 90 videos were used for training, 10 for validation, and 10 for testing. The number of frames in which the diaphragm appears and does not appear in each of these sets is detailed in Table 1. For the training set, the frames without diaphragm appeared in 45/90 videos. For the validation and test sets, the frames without the diaphragm appeared in 6/10 and 4/10 videos respectively.

**Table 1.** Number of frames in each class for diaphragm detection.

|  | Frames with diaphragm | Frames without diaphragm | Total frames |
| --- | --- | --- | --- |
| Train | 3822 (87%) | 565 (13%) | 4387 |
| Validation | 393 (80%) | 100 (20%) | 493 |
| Test | 451 (95%) | 24 (5%) | 475 |

Table 2 shows the sensitivities and specificities and Figure 2 shows the confusion matrices for the PulseNet + SVM model as compared to the benchmark ResNet-50 models. The PulseNet encoder + SVM classifier were both more specific and sensitive in both sets, despite seeing an expected dip in performance on the test set. These results include temporal corrections as previously described. In particular, PulseNet + SVM was much more sensitive than the ResNet models, thereby conserving data. Figure 3 shows a set of predictions, before and after post-processing, as compared to the ground truth, taken from the test set. Common errors in prediction for the PulseNet model were in the transition between the diaphragm disappearing or appearing from view, or when the diaphragm was particularly blurry or unclear.

**Table 2.** Results for diaphragm detection. Sensitivity and specificity are reported with respect to detecting frames with no diaphragm.

|  | Specificity |  | Sensitivity |  | F1 score |  |
| --- | --- | --- | --- | --- | --- | --- |
|  | Validation | Test | Validation | Test | Validation | Test |
| ResNet-50 + CE Loss<br>(benchmark) | 0.96 | 0.97 | 0.5 | 0.42 | 0.86 | 0.94 |
| ResNet-50 + Focal Loss | 0.97 | 0.97 | 0.37 | 0.00 | 0.82 | 0.91 |
| PulseNet + SVM | **0.99** | **0.98** | **0.72** | **0.63** | **0.93** | **0.96** |

**Fig. 2.**
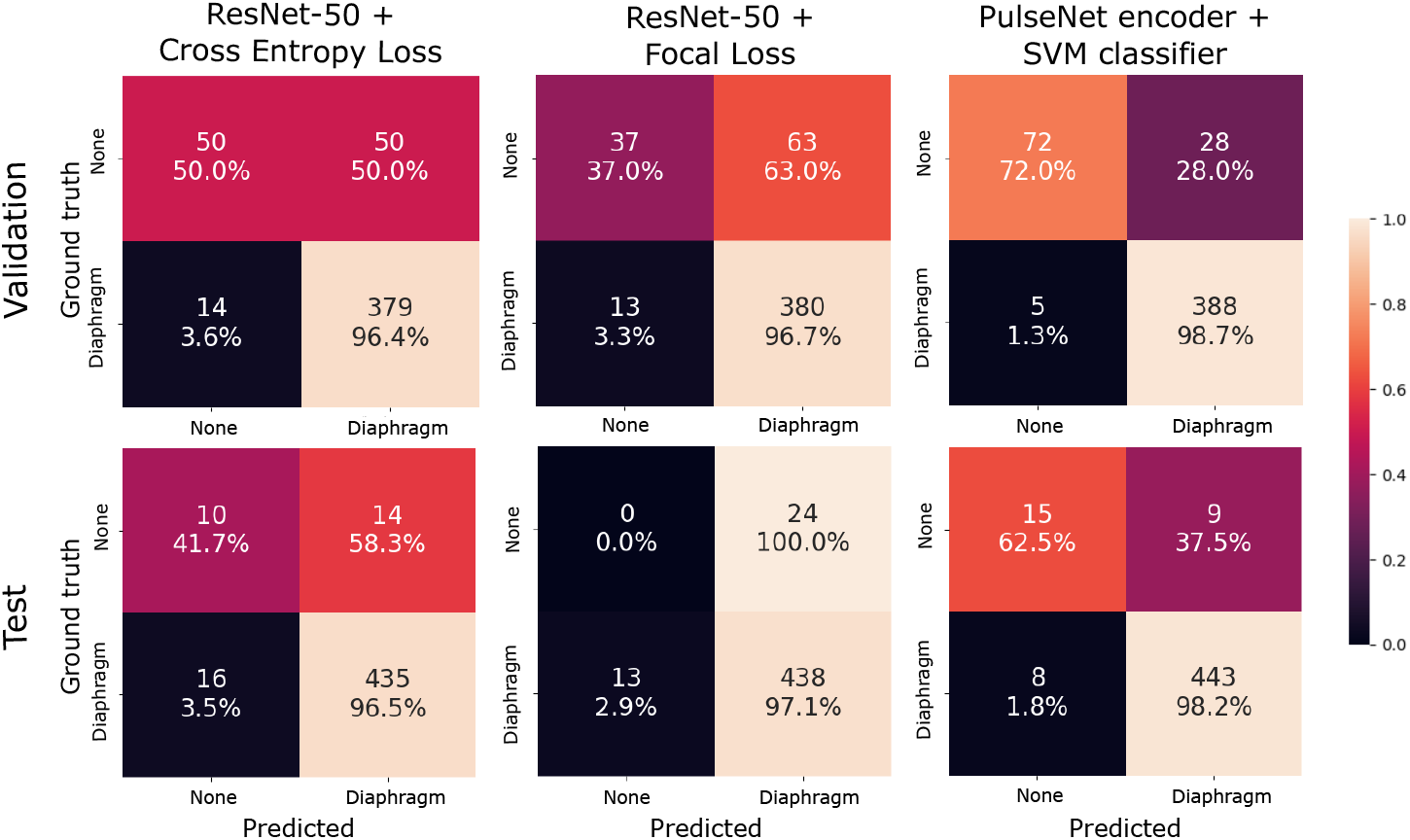
Diaphragm detection confusion matrices.

**Fig. 3.**
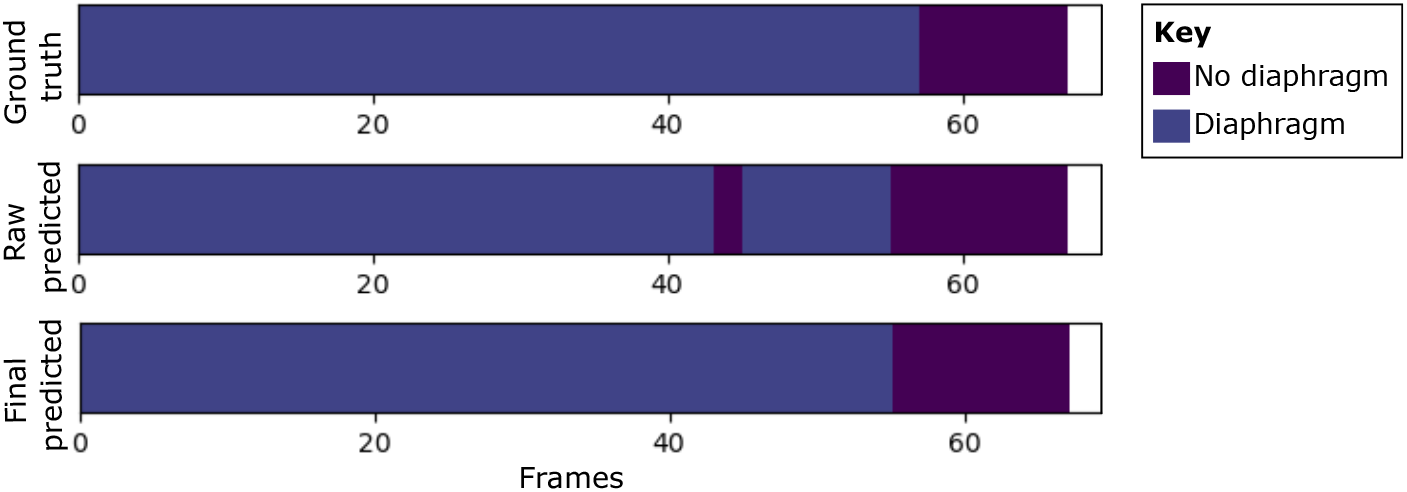
Diaphragm detection for an example video. Frame-by-frame diaphragm detection predictions, both before and after post-processing, compared to the ground truth for an example video with a total number of 66 frames.

### 3.2 Identification of Liver Tissue

There was an improvement (Table 3) in the intersection over union (IoU) when identifying liver tissue using the full pipeline using the diaphragm as an anchor, as compared to directly segmenting liver tissue. The results presented in this table use the SSIM selected ultrasound image to initialise the segmentation model. For comparison, the model was rerun using the first frame of ultrasound video for initialisation; the SSIM method was nearly equivalent to using the first frame with a difference in the mean IoU of <1%. Figure 4 shows examples of failure cases for each model. Figures 4 (a) and (b) show that the direct liver segmentation model incorrectly used vessel walls as the liver tissue boundary. Figure 4 (c) shows an example where both lobes of the liver are in view, and the poor prediction of this tissue by both models.

**Table 3.**
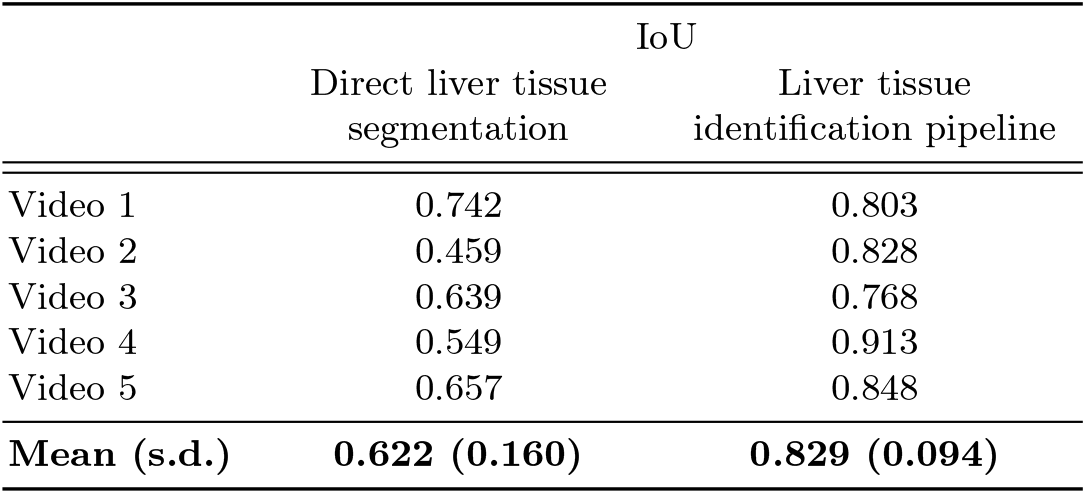
Liver tissue identification results. Comparison in intersection over union (IoU) of directly segmenting liver tissue and inferring liver tissue from diaphragm position. The mean IoU is weighted by the number of frames in each video.

**Fig. 4.**
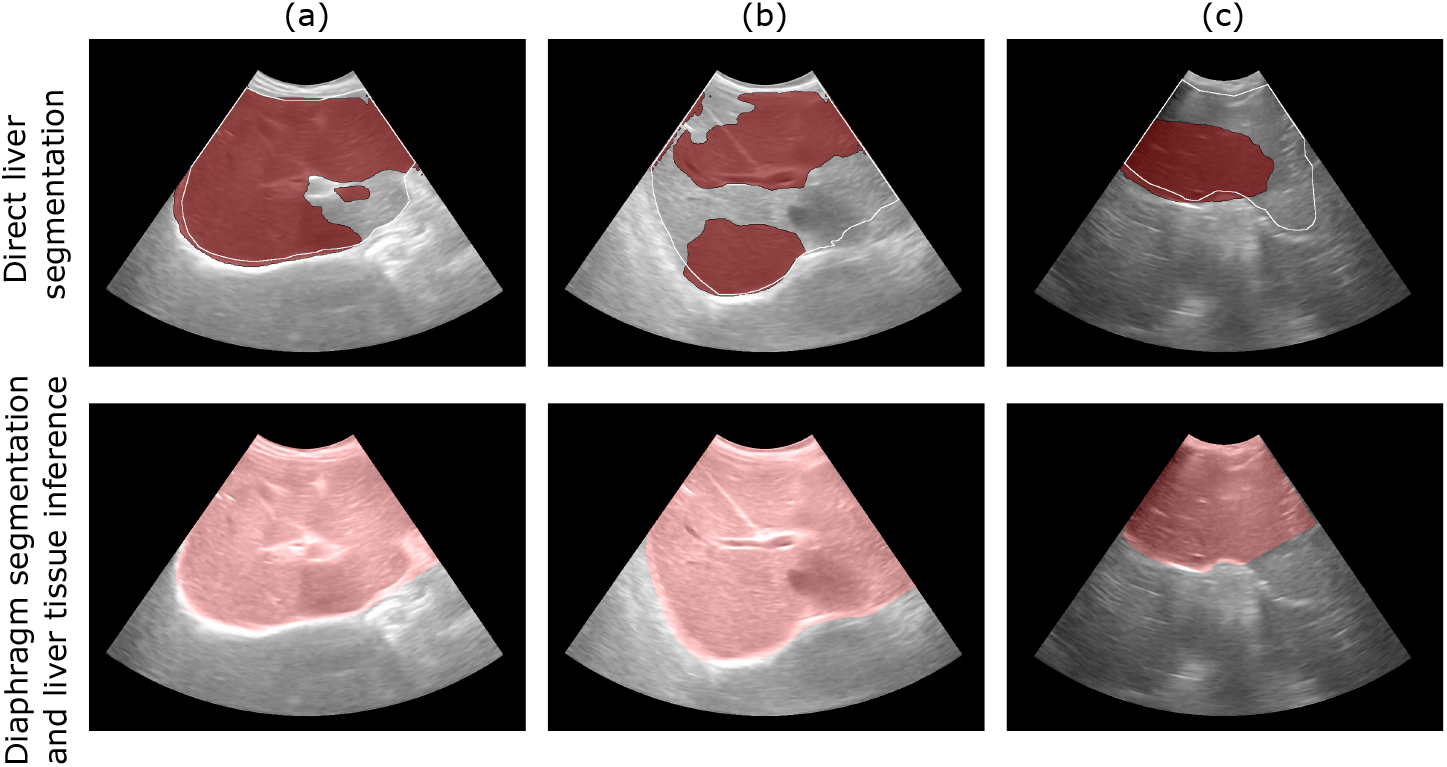
Failure cases. Examples of where the diaphragm segmentation and inference improves liver partitioning.

## 4 Conclusion

A fully automated pipeline for identification of liver tissue in ultrasound video has been presented. This pipeline enables automatic annotation of the diaphragm and liver tissue in large volumes of ultrasound video data, as a processing step before liver disease diagnosis and staging. The liver identification pipeline presented here is more fit-for-purpose than direct liver segmentation. Errors encountered using direct liver segmentation include the detection of other echogenic structures, in particular vessel walls, as the boundary for liver tissue. Not only is this anatomically incorrect, but the results from direct segmentation are not useful as a pre-processing step for schistosomal PPF diagnosis or similarly complex liver diseases requiring whole views of the liver for accurate staging. In particular, schistosomal PPF develops along vessel walls, so using these as a boundary will discard important information for staging. Importantly, the proposed pipeline is suitable for processing data from low-cost, portable ultrasound in rural, resource-limited areas and generalisable to sub-Saharan African populations.

### Limitations

There were several underlying assumptions that underpin this work. Firstly, there could be cases, if free-hand probe sweeps are instead used, where the diaphragm does not appear in an ultrasound video frame but there is liver tissue in that frame. However, this can be avoided by following the same acquisition protocol used in this study. Further to this, the method to infer the position of liver tissue from the diaphragm might provide a crude estimate of liver tissue area in a given frame of ultrasound video, especially when the diaphragm does not stretch fully across the field of view. This issue would be an important consideration if the pipeline were to be used for liver tissue area measurement or volume estimation but does not undermine the purpose of this pipeline; which is to identify the region of interest for liver disease classification models.

### Future work

The generalisability of this pipeline to diseased populations, to children, and to videos with known quality issues should be investigated in future studies. Subsequently, the key next steps for this pipeline include an evaluation as a pre-processing step before the use of a classification model for liver disease staging, including for schistosomal PPF. Importantly, a full pipeline for liver tissue identification and morbidity prediction for diseases like schistosomal PPF could serve as a capacity building tool for trainee sonographers, a risk prediction tool for clinical decision support, and a triage system for screening patients where staff are limited in areas of sub-Saharan Africa.

## Data Availability

Data is not publicly available due to data protection and ethics restrictions related to the ongoing nature of the SchistoTrack Cohort and easily identifiable nature of the participants. Code is available from the authors upon request.

## Ethics Approvals

Data collection and use were reviewed and approved by Oxford Tropical Research Ethics Committee (OxTREC 509-21), Vector Control Division Research Ethics Committee of the Uganda Ministry of Health (VCDREC146), and Uganda National Council of Science and Technology (UNCST HS 1664ES).

## Acknowledgments

We would like to acknowledge Betty Nabatte as the Ugandan project manager; Victor Anguajibi, Timothy Mugume and Benjamin Ntegeka as sonographers for the SchistoTrack study; and the study teams and participants. We would like to acknowledge He Zhao for providing code for the annotation propagation model and advice, and Christopher Ho for assisting with the preparation and storage of ultrasound videos.

ESO receives a DPhil studentship (2593890) associated with the EPSRC CDT in Health Data Science (EP/S02428X/1). GFC receives funding from the Wellcome Trust Institutional Strategic Support Fund (204826/Z/16/Z) and John Fell Fund as part of the SchistoTrack Project, Robertson Foundation Fellowship, and UKRI EPSRC Award (EP/X021793/1). AN acknowledges EPSRC grants EP/X040186/1, ERC grant PULSE (ERC-2015-AdG-694581) and the NIHR Oxford Biomedical Research Centre Imaging Theme. For the purpose of Open Access, the author has applied a CC BY public copyright licence to any Author Accepted Manuscript version arising from this submission.

